# Navigating scarcity: a qualitative study of healthcare workers’ perspectives on essential emergency and critical care in Tanzanian primary health facilities

**DOI:** 10.1101/2024.10.09.24315178

**Authors:** Manji Nyaganya Isack, Dickson Ally Mkoka, Beatrice Mwilike

## Abstract

**Objectives:** To explore the lived experiences of nurses and doctors in delivering essential emergency and critical care (EECC) in Tanzanian primary healthcare (PHC) facilities; to understand the systemic and resource challenges they face; and to document their perspectives on improvements needed for effective EECC service delivery.

**Methods:** A qualitative phenomenological study using in-depth, semi-structured interviews. Data were analysed using reflexive thematic analysis. Three PHC facilities (two health centers and one district-level hospital) in an urban district of the Kilimanjaro Region, Northern Tanzania, were involved. Twelve healthcare workers (six nurses, six doctors) were purposively sampled for their experience in managing critically ill patients. Participants included five men and seven women. The median clinical experience was five years.

**Results:** Thematic analysis produced four themes. (1) A constrained ecosystem of care, defined by severe shortages of space, equipment, drugs, oxygen, and trained personnel. (2) Navigating clinical uncertainty and systemic failure, describing how providers improvised without protocols and relied on a weak, single-ambulance referral system, described as a ‘triage-and-referral’ system. (3) The human cost of systemic failure, leading to preventable deaths and moral distress among providers. (4) Participant-derived blueprint for strengthening EECC, calling for multilevel action: investment in basic resources, human resource development, policy leadership, and facility-level advocacy.

**Conclusions:** PHC workers in Tanzania contend with a system that lacks resources for EECC. They often serve as referral facilitators rather than providers of definitive care. Their experiences provide ground-level evidence to inform the implementation of Tanzania’s 2023-2026 National Strategic Plan on EECC. Strengthening EECC capacity at PHC is paramount to reducing preventable deaths and building a resilient health system.

## Introduction

Critical illness is a significant global health burden and a leading cause of mortality, especially in Sub-Saharan Africa.^1^ About 12.5% of patients in African hospitals are critically ill, with 20.7% dying, of which 20% die within 7 days.^2^ This points to a system failure to provide EECC emphasised by the global community, as ‘the care that all critically ill patients should receive, at a minimum, in all hospitals in the world.^3,4^ This pragmatic, low-cost approach focuses on universal, timely access to first-line treatments, such as oxygen, fluids, and basic monitoring, as an essential component of universal health coverage (UHC) and a strategy to reduce preventable deaths. This shifts the focus from resource-intensive care, feasible only in higher hospitals, to essential care accessible across the entire healthcare system.^5^

In Tanzania and other low-income countries, EECC capacity is severely constrained.^6–9^ Resources are concentrated in urban tertiary hospitals, leaving PHC, the first point of contact for most of the population, greatly under-resourced.^7,8^ Tanzania launched a National Strategic Plan on EECC Services (2023-2026), committing to strengthening service delivery across all levels of care.^10^ While quantitative evidence shows material shortages,^6,7,9^ a critical evidence gap remains: we lack understanding of how nurses and doctors in PHC perceive their ability to deliver EECC, the barriers they face, and their priorities for change.

While our data were collected in 2021, the systemic barriers we document (infrastructure deficits, drug and equipment shortages, lack of protocols, inadequate processes, and unreliable referral systems) are structural problems that evolve slowly in low-resource settings. Evidence over the past decade confirms their persistence.^2,6,7,9,11,12^ The 2025 African Critical Illness Outcomes Study reported that most (68.6%) critically ill patients receive care in general wards without essential treatments.^2^ A 2024 hospital readiness study found that while essential resources were available in hospitals, only 56% were ready for immediate use in wards, with district hospitals scoring below 50%.^9^ Together, these findings indicate that the challenges we identified remain highly relevant. Moreover, our data predate Tanzania’s National Strategic Plan on EECC (2023-2026),^10^ positioning this study as a valuable pre-implementation baseline for evaluating progress under this policy.

This qualitative study aimed to address this gap by exploring the perspectives of nurses and doctors on the delivery of EECC in Tanzanian PHC facilities. We sought to (1) describe their experiences managing critically ill patients, (2) explore the systemic and resource challenges they face, and (3) document their insights into improvements needed, providing ground-level evidence to guide Tanzania’s national EECC strategy and similar efforts in low-resource settings.

## Methods

### Study design

We employed a descriptive phenomenological qualitative design. This approach was most appropriate to gain a detailed, rich understanding of the lived experiences (emotional, practical, systemic, and ethical) of nurses and doctors in providing EECC in Tanzanian PHC facilities.

Building on the objectives outlined above, the study focused on exploring their experiences, perceived challenges, and advocated solutions related to EECC delivery. The study was conducted between May and June 2021 in an urban district in the Kilimanjaro Region, Northern Tanzania.

### Patient and public involvement

None

### Setting and context

Using data from the 2022 census, which reported a regional population of 1,861,934 and an average annual growth rate of 1.3% from 2012 to 2022, we estimate that the 2021 population of the Kilimanjaro Region was approximately 1,838,000.^13^ Purposive sampling was used to select three PHC facilities that represented a spectrum of capacities relevant to EECC delivery. We included one district-level hospital (anonymized as F1) and two health centers (anonymized as F2 and F3). We chose this setting deliberately. Urban PHC facilities in Tanzania typically have greater resource proximity and staffing levels than their rural counterparts. Studying the challenges of EECC delivery in this relatively resourced context provides a conservative, ‘best-case’ analysis, highlighting systemic gaps that are almost certainly more severe in remote settings. In the Tanzanian system, these facilities are tasked with initial patient assessment, stabilization, and management. However, their capacity to manage critical illness is limited; complex cases must be referred to higher-level facilities within the region, specifically Mawenzi Regional Referral Hospital or the Kilimanjaro Christian Medical Centre (KCMC), a tertiary zonal hospital, through the national pyramidal referral system. Studying PHC facilities was crucial because the experiences, challenges, and perspectives of healthcare workers on EECC have not been explored in depth.

### Eligibility criteria

The study involved nurses and doctors from three PHC facilities. Inclusion criteria: (1) at least one year of work experience at the facility; (2) direct recent experience in managing a critically ill patient requiring EECC within the six months before the study; and (3) involvement in referral processes. Exclusion criteria: being on extended leave during the data collection period or holding a purely non-clinical, administrative role. Participants from various units (general wards, outpatient, observation room, theatre, labour ward) were purposively selected, with assistance from facility heads to ensure a diverse range of perspectives. The researcher approached eligible staff in person, explained the study, and invited them to participate.

### Sampling strategy

While phenomenological studies range from 5 to 25 participants,^14^ data collection in this study was guided by thematic saturation.^15^ The aim was to achieve a point where no new information emerged at the level of the shared phenomenon, ‘the experience of providing EECC in PHC facilities’, rather than at the level of individual facilities. Interviews continued until no new themes emerged. After about eight interviews, the central themes related to challenges in EECC, including resource scarcity, referral barriers, and emotional burden, became consistent, indicating initial saturation. Although participants were drawn from different facilities, their reports revealed a common experience of the shared phenomenon, thereby supporting thematic saturation. The final four interviews confirmed and deepened these themes, yielding no new themes. Data collection and analysis were conducted iteratively, although no formal repeat interviews were conducted. All approached eligible healthcare workers agreed to participate (no refusals).

### Researcher characteristics and reflexivity

Before the study, the research team discussed their backgrounds and potential biases. The primary researcher (MNI) is an experienced clinician familiar with the study setting, which provided deep contextual understanding but risked preconceived assumptions about systemic challenges. To mitigate this, the research team adopted a bracketing approach, setting aside their preconceived ideas, biases, and assumptions during data collection and analysis to remain open to the participants’ actual experiences.

### Techniques to enhance trustworthiness

Trustworthiness was pursued using Lincoln and Guba’s criteria.^16–18^ We established credibility through prolonged engagement, iterative data collection until saturation, investigator triangulation (all three authors independently coded and discussed themes), and member checking: following each interview, the first author transcribed the audio recording verbatim. These Kiswahili transcripts were then returned to the participant, either in person during a subsequent facility visit or via email. Participants were given two days to review and provide feedback; the author incorporated the feedback into the final transcripts, which were then translated into English for analysis. This ensured the findings were grounded in participants’ meanings and multiple analytical perspectives.

Dependability was ensured through a detailed audit trail of methodological decisions and a structured codebook, while confirmability was ensured through reflexive journaling of analytical decisions. These made the research process transparent and traceable. The detailed description of the setting enables the transferability of findings to similar settings by informing judgments about underlying methodological decisions. Reporting of this manuscript adhered to the Standards for Reporting Qualitative Research (SRQR) Checklist (Supplementary file 1).^19,20^

### Data collection

A semi-structured interview guide (Supplementary file 2) was developed based on the literature on EECC challenges in low-resource settings. Before piloting via two interviews at a non-participating health centre in the same region, the guide was reviewed for clarity and contextual fit by five clinical experts from similar non-participating hospitals and refined accordingly. MNI conducted the face-to-face interviews in Swahili in private hospital rooms, lasting 25-40 minutes and audio-recorded with no third parties present. Data were collected between May and June 2021. To enhance contextual understanding and support data interpretation, the researcher conducted field observations and reviewed relevant facility documents and records.

### Data analysis

We analysed the data using reflexive thematic analysis with an inductive, interpretive (latent) approach. This data-driven process ensured that themes emerged directly from the interviews. By looking beyond surface-level (semantic) comments, we interpreted the underlying assumptions and ideologies that shaped participants’ experiences, enabling us to understand the broader systemic structures influencing their healthcare behaviour. Braun and Clarke’s six-step framework guided the thematic analysis,^21,22^ and its reporting adhered to the Reflexive Thematic Analysis Reporting Guidelines (RTARG) for coherence and transparency.^23^

The analysis was a collaborative, iterative process involving all three authors. First, they collectively reviewed all transcripts against recordings to correct errors and ensure data accuracy. The 12 transcripts were randomly distributed, with each responsible for the in-depth reading and initial coding of 4 transcripts to support diverse analysis perspectives. Each researcher independently engaged in data familiarization and generated initial codes, identifying meaningful units related to the experiences of providing EECC.

The team then convened for a series of analytical meetings. They presented, compared, and discussed their initial codes, collaborating to develop a unified preliminary codebook. Through further discussion and repeated comparisons with the raw data, we organized codes into candidate themes. These themes were continuously reviewed, refined, merged, and separated over multiple meetings to ensure they coherently captured the richness of the data and accurately reflected the participants’ perspectives. This process continued until we reached consensus on the final thematic structure.

## Results

### Participants characteristics

Interviews were conducted with 12 healthcare workers: 6 nurses and 6 doctors. Participants included five men and seven women. Participants’ ages ranged from 24 to 45 years, with a median of 31; seven were 25 to 35 years, and five were 35 to 45 years. Professional cadres and qualifications were: three Enrolled Nurses (Certificate), four Clinical Officers (Diploma), two Assistant Nursing Officers (Diploma), two Medical Doctors (Bachelor’s degree), and one Nursing Officer (Bachelor’s degree). Clinical experience ranged from 1 to 23 years, with a median of 5 years (interquartile range 3.5 to 13 years). Participants were recruited from the three study facilities: five from the district-level hospital (F1: P01-P05), four from one health centre (F2: P07-P10), and three from the second health centre (F3: P06, P11, P12).

### Interview findings

Thematic analysis yielded four main themes: (1) A constrained ecosystem of care; (2) Navigating clinical uncertainty and systemic failure; (3) The human cost of systemic failure; and (4) Participant-derived blueprint for strengthening EECC. These are accompanied by 15 sub-themes and 56 codes (Table 1). The findings revealed the systemic challenges healthcare workers face in delivering EECC in Tanzanian PHC settings, as well as the solutions they envision. The first three themes detail the experienced reality of scarcity and its consequences, while the fourth encapsulates their collective vision for change. Supporting quotations, tagged with participant and facility identifiers (e.g., P05, F1), illustrate both the universality and the depth of these experiences.

**Table 1.** Final thematic structure: themes, sub-themes, and codes.

| Theme | Context | Sub-Theme | Codes |
| --- | --- | --- | --- |
| 1. A constrained ecosystem of care | The setting | Lack of space and infrastructure | Lack of emergency department (ED) or Intensive Care Unit (ICU); congested observation room; space improvisation (e.g., doctor's room used for emergency care in the absence of a dedicated ED); delayed identification and management of critically ill patients |
|  |  | Critical shortages of utilities and equipment | Lack of monitoring equipment; unreliable oxygen supply; equipment improvisation (e.g., ambu-bag as oxygen substitute, alternating oxygen delivery devices) carry hypoxia risk |
|  |  | Lack of essential drugs and supplies | Frequent stock-out of essential drugs; service gap driving unnecessary referrals |
|  |  | Human resources and deficits in training | Staff shortages; lack of EECC-trained personnel; knowledge and skills gap; gap in curriculum training for EECC; inability to recognize critical illness; inability to prioritize care; lack of shared understanding of critical illness among staff; knowledge gap driving unnecessary referrals |
| 2. Navigating clinical uncertainty and systemic failure | The action | Relying on memory and basic skills | No formal triage system or protocols for critical illness management; reliance on basic skills/memory/clinical observation |
|  |  | Referring as a default response to resource gaps and to manage ethical risk | Referral as default response to resource gaps; referral used to manage ethical risk; facility function shifts from stabilisation to exit facilitation ("triage-and-referral" system) |
|  |  | The ambulance bottleneck | Single ambulance bottleneck causes referral delays; deterioration while waiting for an ambulance; referral journey without oxygen or essential equipment; equipment improvisation (e.g., ambu-bag as oxygen substitute, alternating oxygen delivery devices) carries hypoxia risk; ambulance becomes basic transport, not a treatment unit; patient deterioration or death during referral |
|  |  | When all else fails, improvising to keep patients alive | Improvisation as a necessary skill; equipment improvisation (e.g., ambu-bag as oxygen substitute, alternating oxygen delivery devices) carries hypoxia risk; role improvisation (relying on untrained staff) increases clinical risk; referral pathway bypassing (direct to tertiary hospital) shifts burden upward |
| 3. The human cost of systemic failure | The consequence | Preventable deaths and disabilities | Preventable deaths due to lack of basic EECC (e.g., bleeding control, oxygen); delays leading to long-term disability (e.g., amputation); deaths during referral journey due to lack of initial stabilisation |
|  |  | Moral and emotional distress | Moral distress from inability to provide needed care; guilt and sense of failing patients; overwhelmed by patient load with limited resources; conflict between professional values and what the system allows |
|  |  | The financial and logistical burden on patients and families | Families burdened with obtaining supplies cause treatment delays; prolonged patient suffering due to delays; financial hardship and emotional stress on families; inequity of shifting costs to those least able to afford care |
| 4. Participant-derived blueprint for strengthening EECC | The response | Foundational investment in infrastructure and basic resources | Reliable oxygen; essential drugs and equipment; dedicated space and infrastructure for EECC |
|  |  | Comprehensive human resource development | Training for all staff levels; free accessible courses; mentorship from specialists; curriculum reform |
|  |  | Policy and system-level leadership | Government prioritisation; integration into UHC package; guidelines to standardise care; referral system coordination; affordable EECC |
|  |  | Facility-level advocacy and researcher engagement | Management advocacy; documenting burden; researcher engagement. |

## Theme 1: A constrained ecosystem of care

The foundational experience was characterized by a severe lack of resources, creating an environment in which delivering even basic EECC was structurally impossible. This ecosystem failure manifested in critical shortages of space, equipment, drugs, and skilled personnel.

### 1.1 Lack of space and infrastructure

Participants universally reported the lack of dedicated emergency care spaces, a fundamental requirement for any EECC system. The absence of EDs and ICUs forced care to be provided in inadequate multipurpose rooms. Two participants in F1 described the impact: *“Our observation room is small with only three beds…congested…does not permit proper segregation of patients by age or clinical status…limits emergency responses where you will be forced to evacuate other patients first or shift to another room, thereby losing critical minutes. This was our biggest challenge during the COVID-19 outbreak”* (P02, P04). This illustrates how the physical environment hinders timely intervention, a fundamental component of EECC.

A critical infrastructure gap was the absence of dedicated emergency spaces, particularly in health centres where care was relegated to doctors’ unequipped rooms. As a nurse noted, *“We don’t have an ED/ICU”* (P06, F3), with care happening *“…just here in the doctor’s room”* (P07, P08, F2). This scarcity fundamentally halted flow capacity and systematic patient assessment. Consequently, as one participant described, *“critically ill patients are not identified early in the cloud and [fail to] receive prompt care”* (P10, F2), directly linking the physical environment to delays in the EECC function of triage.

### 1.2 Critical shortages of utilities and equipment

The inability to provide oxygen, a cornerstone of EECC, characterized the breakdown. Reliance on electricity-driven concentrators without backup power turned basic treatment into a risky gamble. A nursing officer in the labor ward explained the systemic flaw: *“We lack oxygen backup cylinders… When I get a patient, and there’s no electricity, the baby won’t receive oxygen on time. I use an empty ambu-bag to resuscitate, until electricity is restored… risking the patient’s survival…”* (P06, F3). This reveals improvisation as a forced competence, in which workers use inadequate tools to fulfill an essential EECC function. Furthermore, the lack of basic monitoring equipment limited objective assessment. A clinical officer in F2 cited: *“We lack a pulse oximeter… leaving reliance on observation”* (P10).

### 1.3 Lack of essential drugs and supplies

The routine stock-outs of essential emergency medications meant the system could not reliably deliver first-line, life-saving drug therapies, a fundamental component of EECC. A common frustration highlighted this failure: *“…sometimes you are out of salbutamol for nebulization… the time is passing by, and I am thinking, come on, how is this patient going to survive?”* (P03, F1). This conveys the helplessness of being a spectator to clinical deterioration due to a missing, low-cost item. Furthermore, the response to this scarcity often exacerbated delays: *“…when I lack essential drugs… I prefer to refer my patient early, instead of sending them to private pharmacies… while delaying care”* (P05, F1), illustrating a rational yet system-damaging coping mechanism in which early referral becomes the default to manage clinical and moral risk.

### 1.4 Human resources and deficits in training

Constraints were both human and material. A critical staff shortage was compounded by a widespread, foundational lack of EECC knowledge and skills among those present, creating a significant gap between the required and available competencies. All participants reported this deficit. The dual challenge was clear: *“…apart from lacking equipment…we lack staff. We are few, and largely, we also lack updated knowledge and skills…”* (P01, F1). This knowledge gap hindered the fundamental principle of triage, with direct clinical consequences.

As a medical doctor reported: *“You could be on shift with someone who does not recognise if a patient is in critical condition. You order, let the drip run faster, but when you come, it is slow. Badly, has left the patient, is busy with an outpatient who just came for a tetanus toxoid injection. You may think it is negligence, but in reality, they don’t know. People should be taught to identify critical illness and prioritize care”* (P02, F1). This points to the absence of a shared, basic understanding of critical illness among frontline staff, a prerequisite for any coordinated EECC response. As the participant added, this deficiency directly drove system strain: *“…if we had staff capable… we wouldn’t need to refer some patients.”*

## Theme 2: Navigating clinical uncertainty and systemic failure

Within this resource-constrained setting, healthcare workers were forced to navigate profound clinical uncertainty. Lacking protocols and facing an under-resourced referral system, they relied on improvised, individual strategies that often revealed and compounded systemic failures.

### 2.1 Relying on memory and basic skills

In the absence of formal triage systems, the essential first step in any EECC pathway, identifying the critically ill, depended on individual clinicians relying on memory and basic skills, using variable and rudimentary methods. Participants described relying on clinical observation alone *(”We don’t have a formal system for categorizing patients… we usually rely on clinical assessment”* (P07, F2) and on foundational, often outdated, personal knowledge. This was evidenced by descriptions such as *“we usually consider vital signs… look at ABCDE”* (P01, F1) and *“I always consider evaluating… Glasgow Coma Scale; it’s based on methods we learned in school and no new”* (P02, F1). This reliance on memory and basic skills highlights the EECC process’s lack of a standardized, systematic approach to prioritization, a critical failure.

### 2.2 Referring as a default response to resource gaps and to manage ethical risk

Faced with severe resource gaps, referral was not just a clinical decision but a default moral and logistical strategy. Participants described referring early to prevent deterioration, to manage cases they knew they could not manage, and to mitigate ethical risk within a system lacking essential resources, thereby making the organisation of referral a primary clinical task rather than definitive care. The reason for referral was a direct result of a lack of EECC capacity: *“I refer a patient early when I cannot meet their clinical needs due to either a lack of oxygen, drugs, or equipment”* (P01, F1). This reflex is based on an honest evaluation of the facility’s role: *“As a primary health facility, we usually have equipment for the ‘initial care’, which is also lacking, so we have to refer regardless of the seriousness of illness*” (P07, F2). This represents a ‘triage-and-referral’ system, in which the facility’s primary function shifted from providing initial stabilization to facilitating exit.

### 2.3 The ambulance bottleneck

The referral pathway itself was a site of failure. A single ambulance serving multiple facilities created a bottleneck, turning referrals into a high-risk period due to delays and insufficient care. Four participants from health centres described a common, fraught experience how sharing one ambulance often led to emergencies being delayed while the ambulance was in use: “The challenge of sharing one ambulance… is awful… If you get an emergency, you call an ambulance, and you find it is in use… the waiting causes deterioration…” (P08, P10, F2; P11, P12, F3). These delays, beyond logistical issues, led to severe patient deterioration and preventable deaths by the time patients reached tertiary hospitals.

The referral journey was medically risky: *“It takes over seven kilometers to reach KCMC, sometimes without oxygen, or the available ones cannot afford a long journey. You delivered twins with birth asphyxia, you found a single cylinder in the ambulance, and one nasal cannula…You tell the driver ‘Run’, then alternate the same nasal cannula if one turns blue…”* (P06, F3). This transformed the ambulance from a mobile treatment unit into a basic transport vehicle, where EECC (like continuous oxygen) is often impossible to provide, leading to further deterioration and preventable deaths.

### 2.4 When all else fails, improvising to keep patients alive

When the system completely lacked essential resources, providers engaged in creative improvisation, using whatever was at hand to keep patients alive, revealing a hidden, high-stakes skill set driven by urgent need. This extended to four critical areas: equipment, personnel, and pathways.

- Improvising with equipment and delivery devices: faced with absent or inadequate tools, providers devised risky workarounds for basic life support that carried a direct risk of hypoxia and ineffective resuscitation. An empty ambu-bag was used to continue resuscitation when backup oxygen cylinders were unavailable during a power outage that turned off the electricity-powered oxygen concentrators. This practice delivers room air (21% oxygen) rather than the near-100% oxygen required to effectively manage hypoxia. During transport, improvisation became even more dangerous, as described by a nurse-midwife managing a twin referral (Sections 1.2 and 2.3; P06, F3 description).
- Improvising roles and using available staff: staff shortages forced reliance on personnel without the necessary skills for critical situations, increasing clinical risk. A participant explained the dilemma during a referral: *“…you leave the labor ward with only a medical attendant who doesn’t have the knowledge or skills to resuscitate the baby or respond to obstetric emergencies…it is a risk”* (P06, F3).
- Improvising referral pathways: at a systemic level, providers developed a workaround for a known problem, the unreliable referral system, bypassing steps in the pyramidal healthcare system (e.g., from health centres directly to tertiary hospitals): *“You send a patient with birth asphyxia or eclampsia to regional hospital; you reach there; they tell you, send to tertiary hospital, we are also sending our patients there… You catch another road… You get frustrated, so next time, instead of roaming with the patient at the district or regional hospital, while deteriorating, I go directly to the tertiary hospital”* (P06, F3). This bypassing, while rational from the provider’s perspective, highlights the disorganization of the referral system and the underdeveloped EECC at lower-level facilities. Significantly, it shifts the entire critical care burden onto tertiary hospitals, thereby undermining the efficiency of the entire health system.

These acts of improvisation, while evidence of frontline resilience, highlight the serious deviation from standard EECC protocols that resource scarcity forces upon providers. They represent not best practices, but last-resort measures to prevent immediate death within a resource-constrained system.

## Theme 3: The human cost of systemic failure

The convergence of resource scarcity and systemic breakdown extracted a heavy human toll, leading to preventable patient harm and psychological distress among healthcare workers, who were acutely aware of the standard care they were unable to provide.

### 3.1 Preventable deaths and disabilities

Participants were clear: preventable patient deaths and disabilities were direct outcomes of system failure to provide EECC. A medical doctor clearly analyzed: *“Many deaths among critically ill patients are preventable, caused by bleeding or lack of oxygen, issues that could be addressed by controlling bleeding or securing airways…”* (P02, F1). He explicitly linked delays to long-term harm: *“Delays can lead to severe outcomes, like amputation.” The referral journey itself added further risk*: *“Not all referred patients reach their destination; some may die en route…critical minutes have been lost without adequate stabilization”* (P12, F3). These are not random tragedies but rather logical, predictable outcomes of a system unable to provide equal EECC services across care levels.

### 3.2 Moral and emotional distress

Healthcare workers described severe moral and emotional distress; the psychological burden of inaction, which conflicted with their professional values and duties. This distress was directly tied to systemic failure: *“This affects my ability to work because if it were something I am capable of doing, but now, because of no equipment, I end up referring the patient. This makes me feel uncomfortable or stressed about my work”* (P09, F2). The emotional burden was heavy: *“… you feel you are failing them because you cannot do anything right…”* (P04, F1) and *“It is challenging to provide adequate, timely care to many patients simultaneously”* (P01, F1). This reveals the human cost of a system with severe resource gaps: the demoralization and burnout of its frontline staff.

### 3.3 The financial and logistical burden on patients and families

When drugs or equipment were unavailable, the burden of obtaining supplies was shifted to families, creating both financial hardship and dangerous delays in care. As one participant explained: *“We have to direct family to buy in outside pharmacies. It takes time, and the patient is suffering”* (P03, F1). These delays worsened clinical deterioration and prolonged suffering. The financial strain on families was severe. A nursing officer summarized the family’s experience as *“Annoying, costly, and stressful”* (P08, F2). A medical doctor framed this as an issue of equity: *“The need for these services will save the poor, who can’t afford costly care in large hospitals”* (P07, F2). Together, these quotes show how system failures shift both the practical work and the financial burden onto those least able to bear them, deepening the inequity of inaccessible care.

## Theme 4: Participant-derived blueprint for strengthening EECC

Confronting these multidimensional challenges, participants consistently urged a multi-level strategy for change. This collective vision constitutes a frontline-derived blueprint for implementing feasible EECC at PHC.

### 4.1 Foundational investment in infrastructure and basic resources

The most urgent need was reliable access to EECC’s fundamental resources: dedicated space, oxygen, drugs, and basic equipment. As participants highlighted: *“I think if we could get a constant supply of backup oxygen cylinders, essential drugs, and equipment, it would greatly impact emergency outcomes”* (P06, F3). Concurrently, *“We wish to work in a well-equipped ED and ICU that allows our patients to get the care they want”* (P03, F1). Generally, the consistent call was for “making the essential, essential”, ensuring the constant availability of low-tech, high-impact resources.

### 4.2 Comprehensive human resource development

Participants identified a foundational gap: the education system fails to equip most frontline health workers with basic knowledge of EECC. They called for curriculum reform, accessible training, and a facility-wide culture of emergency response. As P01, F1 stated, *“Many of us who work here in primary facilities have a certificate or diploma and lack the basics of EECC in our college curriculum. Even those with bachelor’s degrees or higher often don’t have them, as few universities cover them in some cadres. I think it should be included in the curricula of all levels and cadres for a unified understanding”*.

Beyond pre-service education, they emphasized the need for universal, in-service training to create a common understanding: *“Training should be non-discriminatory and should start with watchmen in the gates and medical attendants to all other staff to have a common understanding*” (P07, F2). This vision moves beyond training individuals to building a coordinated, facility-wide EECC capacity.

They critiqued the financial barrier to life-saving courses and pointed to a model for specialist mentorship: *“…many critical care courses such as Basic Life Support and Advanced Trauma Life Support are taught for money… if you can offer frequent free fire extinguisher courses… why can’t you teach trauma for free so that people can save patients?…people are not prioritizing it”* (P02, F1). He added, “*KCMC has many critical care specialists, but are they getting a chance to rotate here and see what we are doing? Or they wait until we bring them, patients? If they still need to work hard now, at the end of the day, I will only connect Ringer’s Lactate or oxygen, or only two stitches to control bleeding, then send…they could have reduced their workload if they could provide training for lower facilities…like how we reduced cesarean section referrals…”* (P02, F1).

### 4.3 Policy and system-level leadership

Participants placed ultimate responsibility for mandating, enabling, and resourcing EECC at the PHC level on national policy and advocated for its integration into the UHC package. Building from successful national-scale program, *“If the government succeeded in scaling down Anti-Retroviral Therapy, essential drugs, vaccines, and family planning, it can also incorporate EECC into the package and facilitate its wide-dissemination”* (P11, F3). This is a direct call for political prioritization and equitable financing to elevate EECC to the status of other essential services. Part of this policy call was to make EECC affordable *“to benefit everyone”* (P07, F2).

### 4.4 Facility-level advocacy and researcher engagement

To close the evidence-action gap, participants called for localized advocacy and the strategic use of research to drive change, implicating both hospital management and researchers as essential agents. They advised, *“hospital management should document how many lives are lost just by the absence of oxygen or staff lacking knowledge”* (P02, F1), *and “send the correct burden report to the government”* (P03, F1*). “You see! It is only a matter of readiness and prioritization”* (P06, F3). Furthermore, they tasked researchers with a translational role, linking evidence to action: *“It is the duty of you who are collecting data to return the clear gap to the management for action”* (P03, F1; P11, F3). This highlights the demand for actionable evidence and views research as a tool for accountability and system reform, embodying the key dissemination principle of ‘returning findings to the source’ to support accurate planning and action.

## Discussion

This qualitative study explored healthcare workers’ perspectives on the delivery of EECC in Tanzanian PHC facilities. It revealed a system with four connected problems: (1) a severely constrained setting lacking basic space, equipment, drugs, and oxygen; (2) reliance on improvised, informal clinical processes and an unreliable referral system that turns transfer into a period of high risk; (3) serious human costs, including preventable patient deaths and disabilities, alongside provider moral and emotional distress; and (4) participant-derived blueprint, calling for investment in basic resources, training reform, national policy leadership, and using research to drive change.

A key strength is the in-depth exploration of lived experiences, providing rich contextual understanding often missing from quantitative readiness surveys. We enhanced trustworthiness through investigator triangulation, member checking, and following SRQR reporting guidelines.^16,17,19^ Purposive sampling of urban PHC facilities provides a focused analysis of challenges in a relatively resourced setting, offering a conservative baseline of system failure. As a qualitative study in a single region, the findings are not generalizable, though they may be relevant to similar PHC facilities. The study focused on nurses and doctors, the cadres most actively involved in bedside EECC delivery, to ensure depth of experiential data; this means the perspectives of other essential staff (e.g., pharmacists, technicians) on system-wide barriers were not captured, and the urban context means that the even greater challenges of rural facilities require separate investigation.

Our findings confirm earlier quantitative evidence of critical care resource gaps in Tanzanian hospitals^7,9^ and align with the highest burden of critical illness in Africa.^2^ We add to this knowledge by moving beyond listing shortages to reveal how the system works and the human costs of scarcity. Our qualitative data explain the “readiness gap” documented recently by Khalid et al (2024).^9^ Our participants described why this gap persists: equipment (e.g., pulse oximeters) was lacking, essential drugs (e.g., salbutamol) were frequently out of stock, oxygen concentrators failed during power outages, trained staff were absent, dedicated ED/ICUs did not exist, and ambulance bottlenecks were common. Similarly, a 2026 study on intra-hospital transport documented infrastructure limitations, resource shortages, and inadequate processes.^11^ Our participants’ experiences of dangerous referral journeys (e.g., ambulances without oxygen, single nasal cannulas for twins) provide a complementary, patient-facing perspective on the same systemic failures. The persistence of these challenges across multiple studies underscores their structural nature and the urgent need for system-level solutions.

We describe the PHC response not simply as “inadequate” but as a rational “triage-and-referral” system where the facility’s primary role shifts from providing definitive EECC to quickly triaging and referring patients to higher-level facilities. The “care” provided becomes logistical (arranging the referral) rather than clinical (resolving the emergency). As a result, early referral becomes a calculated decision by providers to manage clinical and ethical risk, a novel explanation for why referral rates are high.

We also document creative improvisation, the use of makeshift solutions such as oxygen workarounds and informal referral pathways, as a necessary but risky skill. We highlight how frontline knowledge is often marginalized in policy and planning. This is built into the system: when it fails to disseminate essential clinical knowledge, the only safe option for frontline providers is to refer. Therefore, what are often called “unnecessary referrals” result from this flawed system. These two aspects of system failure (improvisation and marginalization of frontline knowledge) are largely missing from standard quantitative facility readiness assessments. Our analysis aligns with the global EECC framework,^4^ detailing specific implementation barriers at the Tanzanian PHC level. The problem is a mismatch between patient needs and system capacity. The national pyramid referral system operates in reverse, with PHC facilitating exit rather than stabilization. The participant-driven blueprint is a practical call to overturn this by enabling high-quality EECC.

For Tanzanian policymakers, this study provides ground-level evidence supporting the priorities of the 2023-2026 National Strategic Plan on EECC.^10^ A recent qualitative study of EECC promotion in Tanzania identified key lessons from policymakers, researchers and senior clinicians, emphasizing collaboration, advocacy, and evidence use.^24^ Our findings complement this top-down policy perspective by documenting the frontline reality: nurses and doctors continue to face infrastructure deficits, drug shortages, and moral distress despite policy commitments. It calls for urgent investment in the basics of EECC (reliable oxygen, electricity, drugs, equipment, personnel, guidelines, system flow), and for integration into pre-service curricula for all cadre levels and the UHC package. It also highlights the hospital managers’ role in documenting burdens and advocating, while researchers act as change agents.

The findings offer insights to guide future studies in similar or unexplored rural settings. Importantly, the urban setting of this study suggests these identified gaps represent a “best-case” analysis; the implications for rural facilities, where distances are longer and resources even scarcer, are likely much worse. This study prompts several critical questions for future research: (1) What is the epidemiology and cost of preventable deaths attributable to EECC failures in Tanzania? (2) How can the participant-derived blueprint be costed, piloted, and integrated into existing PHC strengthening initiatives? (3) What are the specific EECC experiences of healthcare workers in rural PHC facilities, where our urban findings suggest challenges are even greater? A nationwide study that combines surveys and interviews on EECC readiness and provider experiences in urban and rural areas is needed to guide equitable resource allocation under the national strategic plan.

## Conclusion

This qualitative study describes the perspectives of frontline healthcare workers delivering EECC in Tanzanian PHC facilities. It reveals a system in which resource scarcity forces providers to serve as referral facilitators, resulting in preventable patient harm and staff moral distress. The participant-driven blueprint offers a ground-level perspective to inform the implementation of Tanzania’s National Strategic Plan on EECC (2023-2026). Strengthening PHC’s EECC capacity is a fundamental requirement for a resilient, equitable healthcare system that saves lives and supports its workforce.

## Supporting information

Supplimentary File 1The SRQR Checklist

Supplimentary File 2 Semi-Structured Interview Guide

## Acknowledgements

We are grateful to the participants for sharing their time and invaluable experiences. We also thank the administration of the participating facilities for their support.

## Author contributions

MNI, as the guarantor, conceptualized and designed the study, conducted the investigation and data collection, managed and transcribed the data, performed the formal analysis, drafted the original manuscript, and provided project administration. DAM and BM contributed to the study’s conceptualization and design, as well as to formal analysis, validation, and supervision. All authors contributed to data interpretation, manuscript review and editing, and approved the final manuscript.

## Statements and declarations

### Ethical considerations

Ethical approval was granted by the Research and Ethics Committee of Muhimbili University of Health and Allied Sciences on 9 April 2021 (MUHAS-REC-04-2021-552).

## Consent to participate

All participants provided written informed consent before enrollment. For confidentiality, participants are referred to as (P01-P12), study facilities as (F1-F3), and the corresponding district name where the study was conducted is omitted.

## Consent for publication

Not applicable

## Data availability statement

All generated data underlying the findings of this study are presented within the article. The interview transcripts contain potentially identifying information about participants and facilities; therefore, they are not publicly shared to protect confidentiality. The corresponding author holds the data. Queries regarding data access and reasonable requests for de-identified data can be directed to the Chairman of the Muhimbili University of Health and Allied Sciences Research and Ethics Committee.

## Abbreviations

ED: Emergency Department
EECC: Essential Emergency and Critical Care
ICU: Intensive Care Unit
KCMC: Kilimanjaro Christian Medical Centre
PHC: Primary Health Care
UHC: Universal Health Coverage

