## Supplementary material for "Navigating scarcity: a qualitative study of healthcare workers’ perspectives on essential emergency and critical care in Tanzanian primary health facilities": Supplimentary File 1The SRQR Checklist

### The SRQR reporting checklist

For checking that qualitative health research articles can be understood and used by everyone

> ⓘ Note
>
> If you have not used a reporting guideline before, read about how and why to use them and check whether SRQR is the most applicable reporting guideline for your work.
>
> Reporting guidelines are most useful when used early in research. When writing a manuscript or application, consider using the Full Guidance where you'll see explanations and examples for each item.
>
> After writing, demonstrate adherence by completing this checklist:
>
> 1. Specify where each item is described (see Note 1).
> 2. Cite this checklist (See Note 2).
> 3. Include your completed checklist as a supplement when submitting to a journal so that future readers can use it to find information.

| Manuscript: Navigating scarcity: a qualitative study of healthcare workers' perspectives on essential emergency and critical care in Tanzanian primary health facilities | | |
|---|---|---|
| | Item Description | Location (or reason for not reporting) |
| **Title & Abstract** | | |
| Title | Describe the nature and topic of the study. Identify the study as qualitative or indicate the approach or data collection methods. | ✅ **COMPLETE.** "A Qualitative Study" is in the **Title**. |
| Abstract | Summarise the key elements of the study using the abstract format of the intended publication. | ✅ **COMPLETE.** The structured abstract (Objectives, Design, Setting, Participants, Results, Conclusions) meets the *BMJ Open* format. **Location: Abstract**. |
| **Introduction** | | |
| Problem Formulation | Describe the problem/phenomenon studied, its significance, relevant theory and empirical work, and gaps in current knowledge. | ✅ **COMPLETE.** The need for frontline perspectives on EECC in Tanzanian PHC is clearly established. **Location: Introduction, paragraphs 1 and 2**. |
| Purpose or research question | Describe the purpose of the study and specific objectives or questions. | ✅ **COMPLETE.** The three objectives (experience, challenges, perspectives) are explicitly stated. |

| | | |
|---|---|---|
| | | **Location: Introduction, final paragraph**. |
| **Methods** | | |
| Qualitative approach and research paradigm | Describe your qualitative approach, your guiding theory (if appropriate), research paradigm, and reasons for your choices. | ✅ **COMPLETE.** Descriptive phenomenological design is stated and justified. **Location: Methods, "Study design" subsection**. |
| Researcher characteristics and reflexivity | Describe how researchers' characteristics may influence the research, including personal attributes, qualifications/experience, relationship with participants, assumptions, and/or presuppositions; potential or actual interaction between researchers' characteristics and the research questions, approach, methods, results and/or transferability. | ✅ **COMPLETE.** The authors' information is provided. A dedicated reflexivity section details the researcher's positioning and bracketing. **Locations: Author affiliations & information; Methods, "Researcher characteristics and reflexivity" subsection**. |
| Context | Describe the setting/site(s) in which the study was conducted, why it was selected, and any other salient contextual factors that may influence the study. | ✅ **COMPLETE.** Urban PHC context, facility selection, and rationale are clearly described. **Location: Methods, "Setting and Context" subsection**. |
| Sampling strategy | Describe how and why research participants, documents, or events were selected; criteria for deciding when no further sampling was necessary, and the rationale for those criteria. | ✅ **COMPLETE.** Purposive sampling and thematic saturation are well described. **Location: Methods, "Eligibility criteria" and "Sampling strategy" subsections**. |
| Ethical issues pertaining to human subjects | Describe any approval by an appropriate ethics review board and participant consent, or explain any lack thereof. Describe any other confidentiality and data security issues. | ✅ **COMPLETE.** IRB approval, permissions, consent, confidentiality, and data security are reported. **Location: Declarations, "Ethical consideration" and "Data availability statement" subsections, and Methods, "Setting and Context" subsection.** |
| Data collection methods | Describe the types of data collected; details of data collection procedures including (as appropriate) start and stop dates of data collection and analysis, iterative process, triangulation of sources/methods, and modification of procedures in response to evolving study findings. Describe your rationale for these choices. | ✅ **COMPLETE.** Interviews, observations, a timeline, and an iterative process are detailed. **Locations: Methods, "Data collection," and "Study design" subsections.** |
| Data collection instruments and technologies | Describe any instruments (e.g., interview guides, questionnaires) and devices (e.g., audio recorders) used for data collection; describe | ✅ **COMPLETE.** Interview guide development, validating for cultural appropriateness, piloting, and audio |

| | | |
|---|---|---|
| | if/how the instrument(s) changed over the course of the study. | recording are described. **Location: Methods, "Data collection" subsection**. |
| Units of study | Describe the number and relevant characteristics of participants, documents, or events included in the study. Describe the level of participation. | ✅ **COMPLETE.**<br>Participant demographics are detailed in **Results, "Participants Characteristics"**; Table 1.<br><br>The level of participation/roles (inclusion of active frontline staff based on direct recent experience in managing a critically ill patient, involvement in referrals, face-to-face in-depth interviews, interview time (25-40 minutes), and member checking of transcripts) is detailed and justified. **Methods, "Eligibility criteria", "Data collection"** and **"Techniques to enhance trustworthiness"** (for member checking). |
| Data processing | Describe the methods for processing data prior to and during analysis, including transcription, data entry, data management and security, verification of data integrity, data coding, and anonymisation / deidentification of excerpts. | ✅ **COMPLETE.** Transcription, translation, and data security are described. **Locations: Methods, "Data collection" and "Data analysis" subsections**. |
| Data analysis | Describe the process by which inferences, themes, etc. were identified and developed, including the researchers involved in data analysis; usually references a specific paradigm or approach. Describe why you chose this process. | ✅ **COMPLETE.**<br>**What we did**: "Reflexive thematic analysis based on Braun and Clarke's (2006) six-phase framework," **How we did it:** "Collaborative coding by all three authors," and **why we did it this way:** Thematic analysis aligning with our phenomenological aim and preferable than content analysis for our inductive goals, are all detailed.<br>**Location: Methods, "Data analysis" subsection**. |
| Techniques to enhance trustworthiness | Describe any techniques to enhance trustworthiness and credibility of data analysis (e.g., member checking, triangulation, audit trail). Describe why you chose these techniques. | ✅ **COMPLETE.** Investigator triangulation, member checking, reflexive journaling, and audit trail, along with their justifications, are explicitly stated. **Location: Methods, "Techniques to enhance trustworthiness" subsection**. |

| Results | | |
|---|---|---|
| Synthesis and interpretation | Describe the main findings (e.g., interpretations, inferences, and themes); might include development of a theory or model, or integration with prior research or theory. | ✅ **COMPLETE.** The four main themes are clearly presented and interpreted. **Location: Results, "Interview findings" paragraph and Table 1**. |
| Links to empirical data | Provide evidence (e.g., quotes, field notes, text excerpts, photographs) to substantiate analytic findings. | ✅ **COMPLETE.** Numerous illustrative quotes with participant IDs are provided for each sub-theme. **Location: Results, Themes 1-4**. |
| **Discussion** | | |
| Integration with prior work, implications, transferability, and contribution(s) to the field | Summarize the main findings, explain how findings and conclusions connect to, support, elaborate on, or challenge conclusions of earlier scholarship; discuss the scope of application/generalizability; identify unique contribution(s) to scholarship in a discipline or field. | ✅ **COMPLETE.** The discussion integrates findings with prior studies and the Tanzanian national plan, discussing contributions and implications. **Location: Discussion, paragraphs 1-5**. |
| Limitations | Discuss the trustworthiness and limitations of findings | ✅ **COMPLETE.** Limitations regarding generalizability, cadre focus, and urban setting are addressed. **Location: Discussion, paragraph 2 and Strengths and Limitations of this study box.** |
| **Other** | | |
| Conflicts of interest | Describe any potential sources of influence or perceived influence on study conduct and conclusions. Describe how these were managed. | ✅ **COMPLETE.** "None declared" is stated. **Location: Declarations, "Competing interests"**. |
| Funding | Describe sources of funding and other support. Describe the role of funders in data collection, interpretation, and reporting. | ✅ **COMPLETE.** "No specific funding" is declared. **Location: Declarations, "Funding statement"**. |

#### 1 How to specify where content is

Tell the reader where they can find information. E.g.,

- Results; paragraph 2
- Methods, Participants; paragraphs 1 & 2.
- Table 3
- Supplement B, para. 4

If you have chosen not to describe an item, explain why. You can do this in the checklist, or as a note below it.

You can describe items in the article body, or in tables, figures, or supplementary materials, and should prioritize items you feel are most important to your intended audience. The order of items in your manuscript does not need to match the order of items in this checklist. You can decide how best to structure your work.

#### 2 How to cite

Describe how you used SRQR at the end of your Methods section, referencing the resources you used e.g.,

> ‘We used the SRQR reporting guideline(1) to draft this manuscript, and the SRQR reporting checklist(2) when editing, included in supplement A’

If you use a reporting checklist, remember to include it as a supplement when publishing so that readers can easily find information and see how you have interpreted the guidance.

1. O’Brien BC, Harris IB, Beckman TJ, Reed DA, Cook DA. Standards for reporting qualitative research: A synthesis of recommendations. Academic Medicine [Internet]. 2014 Sep;89(9):1245–51. Available from: https://journals.lww.com/academicmedicine/fulltext/2014/09000/Standards_for_Reporting_Qualitative_Research__A.21.aspx

2. O’Brien BC, Harris IB, Beckman TJ, Reed DA, Cook DA. The SRQR reporting checklist. In: Harwood J, Albury C, Beyer J de, Schlüssel M, Collins G, editors. The EQUATOR network reporting guideline platform [Internet]. The UK EQUATOR Centre; 2025. Available from: https:/resources.equator-network.org/reporting-guidelines/srqr/srqr-checklist.docx
