## Supplementary material for "Navigating scarcity: a qualitative study of healthcare workers’ perspectives on essential emergency and critical care in Tanzanian primary health facilities": Supplimentary File 2 Semi-Structured Interview Guide

**Supplementary File 2: Semi-Structured Interview Guide**

**Manuscript: Navigating scarcity: a qualitative study of healthcare workers' perspectives on essential emergency and critical care in Tanzanian primary health facilities**

| PART A: Semi-Structured Interview Guide: Exploring Experiences of Essential Emergency and Critical Care (EECC) in Primary Healthcare Facilities—ENGLISH VERSION |
|---|
| **Introduction to Topic: We are interested in understanding your experiences and perspectives on providing care for critically ill patients at this facility, particularly in the context of essential emergency and critical care (EECC).** |
| Section 1: Experiences and Current Practices |
| 1. **Opening Narrative:** Thinking about your work here, how would you describe your overall experience of caring for a critically ill patient?<br>○ Probe: Could you walk me through a recent, specific example from start to finish? What was that process like for you and the patient? |
| 2. **Clinical Processes:** Can you describe for me the typical process for identifying, prioritizing, and managing a critically ill patient here?<br>○ Probe: What happens in a case that becomes too complex for this facility? Can you describe the referral process step by step? Like specific cases you encountered recently and how you handled them? |
| 3. **Service Assessment:** Based on your experience, what is your view on the state of essential emergency and critical care (EECC) services in this setting?<br>○ Probe: What makes you say that? Could you give me a concrete example that illustrates this? |
| Section 2: Challenges and Impacts |
| 4. **Identifying Challenges:** What are the most significant challenges you face when trying to provide good emergency or critical care here?<br><br>**Probes (to be used if not raised spontaneously):**<br>○ **Resources:** Tell me about the availability of things like emergency drugs, oxygen, monitoring equipment, or dedicated space.<br>○ **Systems and Referrals:** How does the referral system work in practice? What makes it difficult?<br>○ **Knowledge and Training:** Do you feel your training has prepared you well for these cases? What ongoing support or training is available? |

| |
|---|
| ○ **Staffing and Teamwork:** Can you describe the staffing situation during an emergency? How does the team coordinate? |
| 5. **Consequences of Challenges:** In your view, how do the challenges you've mentioned affect the care patients finally receive?<br>○ Probe: Do you feel these challenges ever contribute to worse outcomes for patients? Can you explain how? |
| Section 3: Perspectives on Improvement and The Future Role of Primary Health Care |
| 6. **Solutions and Priorities:** If you could change three things to improve care for critically ill patients here, what would they be and why?<br>○ Probe (if needed): Where should we start—with equipment, training, systems, management, or something else? |
| 7. **Training Needs:** Specifically, regarding training, what areas do you think would most significantly improve your ability to manage critically ill patients? |
| 8. **Systemic Role:** Looking at the whole healthcare system, what do you believe should be the core role of a primary health facility like this one in emergency and critical care?<br>○ Probe: And what would need to happen for this facility to fulfill that role? |
| **Closing** |
| 9. **Final Reflection:** Is there anything crucial about your experience providing this type of care that we haven't discussed, or any final thoughts you'd like to share? |

| **Sehemu B: Mwongozo wa Mahojiano: Kuchunguza Uzoefu wa Huduma Muhimu za Dharura na Uangalizi Maalumu kwa Walio Mahututi katika Vituo vya Afya Ngazi ya Msingi— TOLEO LA KISWAHILI** |
|---|
| **Utangulizi wa Mada:** Tunavutiwa na kuelewa uzoefu wako na mtazamo wako kuhusu kutoa huduma kwa wagonjwa walio katika hali mbaya katika kituo hiki, hasa katika muktadha wa huduma muhimu za dharura na uangalizi maalumu (Essential Emergency and Critical Care - EECC). |
| Sehemu ya 1: Uzoefu na Jinsi Huduma Inavyotolewa kwa Sasa |
| **1. Kufungua Mahojiano:** Ukifikiria kuhusu kazi yako hapa, unaelezaje uzoefu wako wa jumla wa kumuhudumia mgonjwa aliekatika hali mbaya “mahututi”?<br>○ **Udadisi:** Unaweza kunipitisha kwenye mfano mahususi na wa hivi karibuni, kutoka mwanzo hadi mwisho? Mchakato mzima ulikuwaje kwako na kwa mgonjwa? |

| |
|---|
| 2. **Utoaji wa Huduma:** Unaweza kunielezea utaratibu wa kawaida wa kutambua, kutoa kipaumbele, na kutoa huduma kwa mgonjwa aliye katika hali mbaya "mahututi" hapa?<br>○ Udadisi: Nini hufanyika kama hali ya mgonjwa ni ngumu/haiwezi kutibika katika kituo hiki? Unaweza kuelezea hatua kwa hatua mchakato wa rufaa kumpeleka mgonjwa hospitali nyingine? Kama mifano mahususi uliyokutana nayo hivi karibuni na jinsi ulivyoshughulika nayo?" |
| 3. **Tathmini ya Huduma:** Kulingana na uzoefu wako, maoni yako ni yapi kuhusu hali ya huduma muhimu za dharura na uangalizi maalumu kwa wagonjwa mahututi (EECC) katika mazingira haya?<br>○ Udadisi: Ni nini kinachokufanya useme hivyo? Unaweza kunipa mfano halisi inayoonyesha hili? |
| **Sehemu ya 2: Changamoto na Athari** |
| 4. **Kutambua Changamoto:** Ni changamoto gani kubwa zaidi unakumbana nazo wakati unapojaribu kutoa huduma nzuri ya dharura au uangalizi maalumu kwa wagonjwa mahututi?<br><br>**Maswali ya Udadisi (kutumika ikiwa haijasemwa moja kwa moja):**<br>○ **Rasilimali:** Nielezee kuhusu upatikanaji wa vitu kama vile dawa za dharura, oksijeni, vifaa vya kufuatilia hali ya mgonjwa, au sehemu/vitengo/vyumba maalumu vilivyotengwa kwa ajili ya kutolewa huduma za dharura au uangalizi maalumu kwa walio mahututi.<br>○ **Mifumo na Rufaa:** Mfumo mzima wa rufaa kumpeleka mgonjwa hospitali nyingine unafanywa kazi vipi? Ni nini kinakuwa ngumu?"<br>○ **Ujuzi na Mafunzo**: Unahisi mafunzo yako yamekutayarisha vyema kwa aina hizi za wagonjwa? Ni usaidizi gani au mafunzo endelevu yanapatikana hapa?<br>○ **Hali ya Uwepo wa Wafanyakazi na Ushirikiano:** Unaweza kuelezea hali ya wafanyakazi wakati inapotokea dharura kwa mgonjwa labda hali imebadilika na anahitaji huduma ya haraka? Timu inashirikiana vipi? |
| 5. **Matokeo ya Changamoto:** Kwa maoni yako, changamoto hizi ulizozitaja zinaathiri vipi huduma ambayo wagonjwa wanapata hatimaye?<br>○ Udadisi: Unahisi changamoto hizi zimewahi/zinaweza kuchangia matokeo mabaya kwa wagonjwa? Unaweza kuelezea vipi? |
| **Sehemu ya 3: Mtazamo juu ya Uboreshaji na Jukumu la Baadaye la Ngazi ya Msingi ya Utoaji Huduma za Afya** |
| 6. **Suluhisho na Vipaumbele:** Kama ungeweza kubadilisha mambo matatu kuboresha huduma kwa wagonjwa walio katika hali mbaya "mahututi" hapa, yangekuwa yapi na kwa nini? |

| |
| --- |
| ○ Udadisi zaidi (ikiwa itahitajika): Tuanzie wapi—kwa vifaa, mafunzo, mifumo, usimamizi, au kitu kingine? |
| 7. **Mahitaji ya Mafunzo:** Hususani, kuhusu mafunzo, ni maeneo gani unafikiria yangeboresha zaidi uwezo wako wa kuhudumia wagonjwa mahututi? |
| 8. **Jukumu Mfumo:** Ukiangalia mfumo mzima wa afya, unaamini nini linaweza kuwa jukumu mama la kituo kama hiki cha utoaji huduma Ngazi ya msingi kwenye utoaji wa huduma za dharura na uangalizi maalumu kwa wagonjwa mahututi?<br>Udadisi: Na nini kitapaswa kutokea/kufanyika ili kituo hiki kitimize jukumu hilo? |
| **Kufunga Mahojiano** |
| 9. **Fikra za Mwisho:** Kuna jambo lolote muhimu kuhusu uzoefu wako wa kutoa aina hii ya huduma ambalo hatujajadili, au mawazo yoyote ya mwisho ungependa kupeleka ujumbe? |
